# Vaccine effectiveness against SARS-CoV-2 infection and COVID-19-related hospitalization with the Alpha, Delta and Omicron SARS-CoV-2 variants: a nationwide Danish cohort study

**DOI:** 10.1101/2022.04.20.22274061

**Authors:** Mie Agermose Gram, Hanne-Dorthe Emborg, Astrid Blicher Schelde, Nikolaj Ulrik Friis, Katrine Finderup Nielsen, Ida Rask Moustsen-Helms, Rebecca Legarth, Janni Uyen Hoa Lam, Manon Chaine, Aisha Zahoor Malik, Morten Rasmussen, Jannik Fonager, Raphael Niklaus Sieber, Marc Stegger, Steen Ethelberg, Palle Valentiner-Branth, Christian Holm Hansen

**Affiliations:** Department of Infectious Disease Epidemiology and Prevention, Statens Serum Institut, Copenhagen, Denmark; Division of Infectious Disease Preparedness, Data Integration and Analysis, Statens Serum Institut, Copenhagen, Denmark; Department of Virus Research and Development Laboratory, Virus and Microbiological Special Diagnostics, Statens Serum Institut, Copenhagen, Denmark; Department of Bacteria, Parasites, and Fungi, Statens Serum Institut, Copenhagen, Denmark; Department of Public Health, Global Health Section, University of Copenhagen, Copenhagen, Denmark

**Author notes:** Corresponding author (MIAG).

## Abstract

**Background:** The continued occurrence of more contagious SARS-CoV-2 variants and waning immunity over time require ongoing re-evaluation of the vaccine effectiveness (VE). This study aimed to estimate the effectiveness in two age groups (12-59 and 60 years or above) of two and three vaccine doses (BNT162b2 mRNA or mRNA-1273 vaccine) by time since vaccination against SARS-CoV-2 infection and COVID-19-related hospitalization in an Alpha, Delta and Omicron dominated period.

**Methods:** A Danish nationwide cohort study design was used to estimate VE against SARS-CoV-2 infection and COVID-19-related hospitalization with the Alpha, Delta and Omicron variants. Information was obtained from nationwide registries and linked using a unique personal identification number. The study included all residents in Denmark aged 12 years or above (18 years or above for the analysis of three doses) in the Alpha (February 20 to June 15, 2021), Delta (July 4 to November 20, 2021) and Omicron (December 21, 2021 to January 31, 2022) dominated periods. VE estimates including 95% confidence intervals (CIs) were calculated using Cox proportional hazard regression models with adjustments for age, sex and geographical region. Vaccination status was included as a time-varying exposure.

**Findings:** In the oldest age group, VE against infection after two doses was 91.0% (95% CI: 88.5; 92.9) for the Alpha variant, 82.2% (95% CI: 75.3; 87.1) for the Delta variant and 39.9% (95% CI: 26.4; 50.9) for the Omicron variant 14-30 days since vaccination. The VE waned over time and was 71.5% (95% CI: 54.7; 82.8), 49.8% (95% CI: 46.5; 52.8) and 4.7% (95% CI: 0.2; 8.9) >120 days since vaccination against the three variants, respectively. Higher estimates were observed after the third dose with VE estimates against infection of 86.0% (Delta, 95% CI: 83.3; 88.3) and 57.6% (Omicron, 95% CI: 55.8; 59.4) 14-30 days since vaccination. Among both age groups, VE against COVID-19-related hospitalization 14-30 days since vaccination with two or three doses was 94.8% or above for the Alpha and Delta variants, whereas among the youngest age group, VE estimates against the Omicron variant after two and three doses were 62.4% (95% CI: 46.3; 73.6) and 89.8% (95% CI: 87.9; 91.3), respectively.

**Conclusions:** Two vaccine doses provided high protection against SARS-CoV-2 infection and COVID-19-related hospitalization with the Alpha and Delta variants with protection waning over time. Two vaccine doses provided only limited protection against SARS-CoV-2 infection and COVID-19-related hospitalization with the Omicron variant. The third vaccine dose substantially increased the protection against Delta and Omicron.

## Background

Mass vaccination of the population is a key strategy to manage the coronavirus disease 2019 (COVID-19) pandemic. However, breakthrough severe acute respiratory syndrome coronavirus 2 (SARS-CoV-2) infections in vaccinated individuals still present a public health challenge (1-3). Multiple studies have assessed COVID-19 vaccine effectiveness (VE) against SARS-CoV-2 infection and severe COVID-19-related outcomes (4-10). A systematic review and meta-regression demonstrated that the VE against SARS-CoV-2 infection and symptomatic disease decreased more than against severe disease six months after two doses (9). However, all the included studies were carried out before the circulation of the Omicron variant (9). The SARS-CoV-2 variants B.1.1.7 (Alpha), B.1.617.2 (Delta) and B.1.1.529 (Omicron) caused rapid increase of SARS-CoV-2 infections worldwide (11) and were classified as variants of concern (VOC) by the World Health Organization (12). Continued emergence of new variants and waning immunity by time since vaccination (3) requires ongoing evaluation of the VE to inform future vaccination strategies. A Danish preprint study has estimated the protection of COVID-19 mRNA vaccines against infection or hospitalization with the Omicron variant and observed relatively poor protection against infection but high VE against COVID-19-related hospitalization after the third dose (13). However, previous studies have observed differences in the VE against SARS-CoV-2 infection with the Alpha, Delta and Omicron variants (4-10) and only few large-scale studies have compared VE against all three variants (10). Despite variation between the variants, a previous study of the risk of hospitalization and death associated with the Delta and Omicron variants observed a significant variation with age (14). The aim of this study was to estimate the effectiveness of two and three doses of the BNT162b2 mRNA (Pfizer/BioNTech) or mRNA-1273 (Moderna) vaccine against SARS-CoV-2 infection and COVID-19-related hospitalization in an Alpha, Delta and Omicron dominant period by time since vaccination in two age groups (12-59 years and 60 years or above).

## Methods

### Study design and setting

We conducted a nationwide cohort study in Denmark. All residents in Denmark are registered in the Danish Civil Registration System (CRS) with a unique personal identification number (CPR number), which is used in all national registries, enabling individual-level linkage between registries (15).

#### The Danish COVID-19 vaccination program

The rollout of COVID-19 vaccines in Denmark was initiated on December 27, 2020. The BNT162b2 mRNA vaccine from Pfizer/BioNTech and the mRNA-1273 vaccine from Moderna are part of the Danish vaccination program. In Denmark, all residents aged 5 years or above are offered two vaccine doses, and those aged 18 years or above are offered a third vaccine dose 4.5 months after the second dose. Denmark has continuously received vaccines during the COVID-19 vaccination rollout. However, relatively few vaccines were available in the initial phase. Hence, the Danish Health Authority determined the order in which population groups were offered vaccination. The populations initially prioritized for COVID-19 vaccination were the most vulnerable citizens and frontline healthcare workers whereas the younger population was invited later (16).

#### SARS-CoV-2 testing

One of Denmark’s main strategies for handling the COVID-19 epidemic was mass testing including unlimited access to free-of-charge SARS-CoV-2 RT-PCR tests at either community testing facilities or hospitals, and rapid antigen tests at a community level. As part of the re-opening of Denmark, a recent negative RT-PCR or rapid antigen test was required for unvaccinated individuals to access indoor public facilities (from March 1 to October 1, 2021, and again from November 11, 2021 to February 1, 2022). These initiatives have ensured a high testing rate for SARS-CoV-2, and the rate of RT-PCR testing in the Danish population is among the highest in the world (11). During the entire study period, it was recommended that a positive rapid antigen test was verified by RT-PCR.

### Study population

The study population included all residents in Denmark aged 12 years or above (18 years or above for three doses) in an Alpha, Delta and an Omicron dominant period. Individuals aged 5-11 years were offered vaccination later and vaccinated with a smaller dose than individuals aged 12 years or above (19). Therefore, we did not included individuals younger than 12 years in the study. Individuals vaccinated with other COVID-19 vaccines than BNT162b2 mRNA or mRNA-1273 were censored at the time of vaccination. Individuals with a reverse transcription polymerase chain reaction (RT-PCR)-confirmed SARS-CoV-2 infection before the start of the study periods and those without any tests during the three variant periods were excluded from the analyses. Only the first positive PCR-test was included.

### Assessment of exposure

All administered COVID-19 vaccines are registered in the Danish Vaccination Registry (DVR) on an individual level, identified by the CPR number (17). Information on the date of vaccine administration and name of the vaccine product was retrieved from the DVR (17).

### Assessment of outcomes (SARS-CoV-2 infection and COVID-19-related hospitalization)

The Danish Microbiology Database (MiBa) receives, in real-time, copies of all laboratory test results from all clinical microbiology departments in Denmark (18). During the COVID-19 pandemic, community-testing facilities were established across the nation, and in early 2021, it became mandatory for private vendors performing SARS-CoV-2 testing to report electronically to MiBa (18). Data on all positive laboratory-confirmed RT-PCR tests were extracted from MiBa (18). Information on rapid antigen tests was not included in this study due to moderate sensitivity in asymptomatic patients compared with RT-PCR (19).

All hospitalizations are registered in an individually identifiable format in the Danish National Patient Registry with date of admission and discharge as well as diagnoses coded according to the International Classification of Diseases, 10th revision (ICD-10) (20). A COVID-19-related hospital admission was defined as any new admission lasting at least 12 hours and occurring between two days before and 14 days after the sample date where SARS-CoV-2 infection with either the Alpha, Delta or Omicron variant was detected.

### Covariates

Information on age (5-years-intervals), sex (male/female) and geographical region (Capital Region of Denmark/Central Denmark Region/Northern Denmark Region/Region Zealand/Region of Southern Denmark) was included as covariates for the association between COVID-19 vaccination and SARS-CoV-2 infection or COVID-19-related hospitalization. Information on all covariates was extracted from the CRS registry (15).

### Statistical analysis

Characteristics of the study population were described using proportions and stratified by vaccination status. Time until SARS-CoV-2 infection (both asymptomatic and symptomatic) or COVID-19-related hospitalization were analyzed in three periods where the relevant variant accounted for at least 75% of all whole-genome sequenced (WGS) RT-PCR confirmed cases: Alpha (February 20 to June 15, 2021), Delta (July 4 to November 20, 2021) and Omicron (December 21, 2021 to January 31, 2022) (Fig 1).

**Fig 1.**
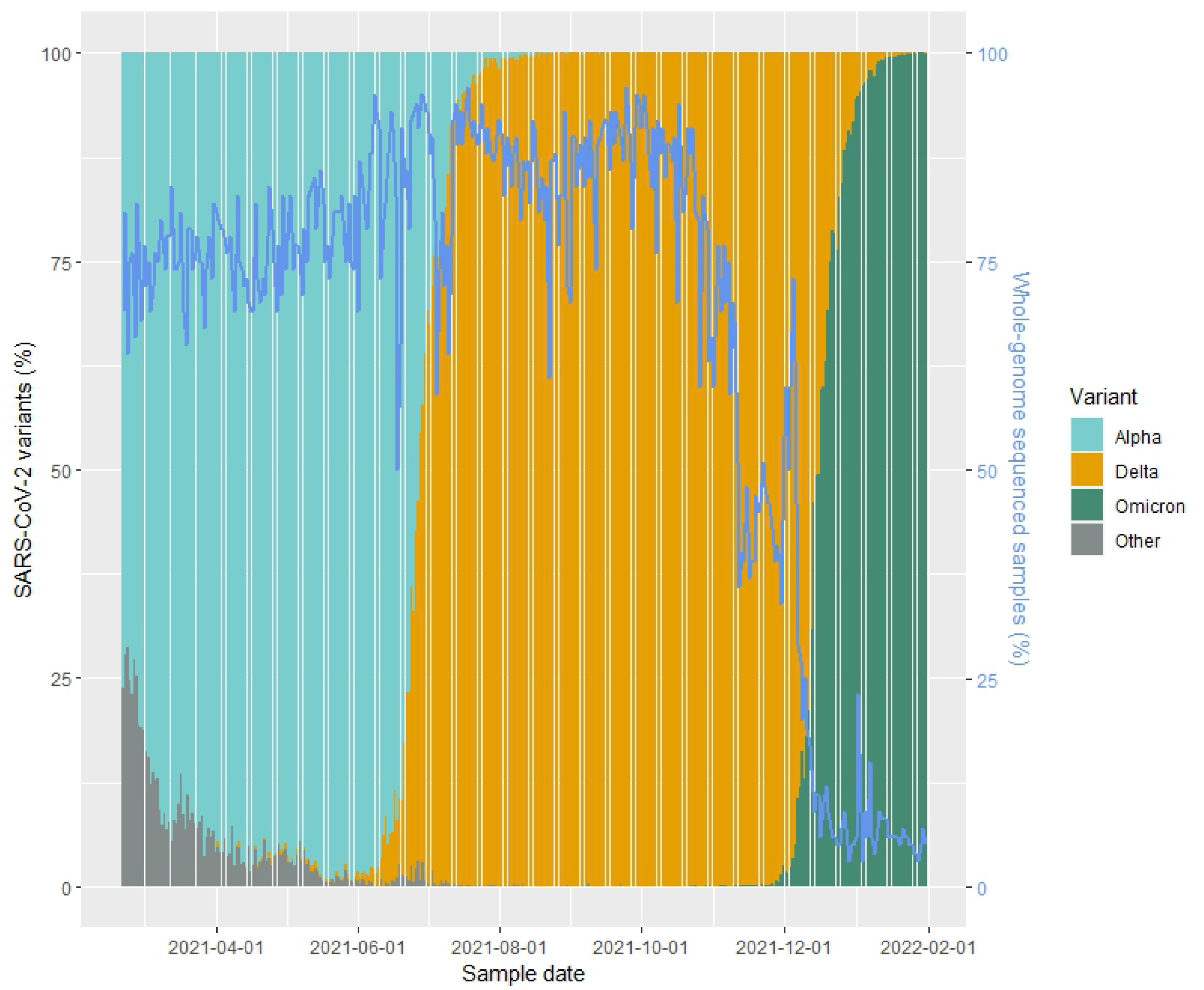
Percentage of whole-genome sequenced samples and the distribution of SARS-CoV-2 variants by sample date.

Separate models were fitted to estimate the VE for the two outcomes, SARS-CoV-2 infection and COVID-19-related hospitalization, and separately for the analysis of two and three doses. Event rates in the vaccinated and unvaccinated exposure groups were compared using hazard ratios estimated in a Cox regression model adjusted for age group, sex and geographical region, with calendar time as the underlying time scale to control for temporal variations in the infection rate. VE was estimated as 1 minus the hazard ratio.

Vaccinated individuals were followed from the start of the study or the date of assumed protection after the second or third vaccine dose, i.e. 14 days since vaccination. All individuals remained in follow-up until the date of SARS-CoV-2 infection, booster vaccination (third dose), death, emigration or the end of study period, whichever occurred first. Unvaccinated individuals remained in follow-up from the start of the study and until the date of their first vaccination, death, emigration, SARS-CoV-2 infection or end of the study, whichever occurred first.

Exposure status was categorized as either unvaccinated or vaccinated with the last dose administered in the past 14-30 days, 31-60 days, 61-90 days, 91-120 days or >120 days. Time falling outside of these categories was not included in the analysis. Data were analyzed using SAS version 9.4.

### Ethical considerations

According to Danish law, ethical approval is not required for anonymized aggregated register-based studies. The study adheres to the Strengthening the Reporting of Observational Studies in Epidemiology (STROBE) (Table S1 STROBE checklist) (21).

## Results

In the Delta dominant period, fewer individuals aged 60 years or above were unvaccinated compared to the Alpha dominant period. In the Omicron dominant period, the majority in both age groups was vaccinated with two or three doses. In the Alpha and Delta dominant period, the median age within each age group was slightly lower in the unvaccinated individuals compared to individuals vaccinated with two or three doses. Across all periods and age groups, the majority of the study population was vaccinated with the BNT162b2 mRNA vaccine (84.9-98.0%) (Table 1).

**Table 1.** Characteristics of the study population by vaccination status.

|  | **Alpha**<br>February 20 – June 15, 2021 |  | **Delta**<br>July 4– November 20, 2021 |  |  |  |  |  | **Omicron**<br>December 21, 2021 – January 31, 2022 |  |  |  |  |  |
| --- | --- | --- | --- | --- | --- | --- | --- | --- | --- | --- | --- | --- | --- | --- |
|  | **60 years or above** |  | **12-59 years** |  |  | **60 years or above** |  |  | **12-59 years** |  |  | **60 years or above** |  |  |
|  | **Unvaccinated** | **Two doses** | **Unvaccinated** | **Two doses** | **Three doses** | **Unvaccinated** | **Two doses** | **Three doses** | **Unvaccinated** | **Two doses** | **Three doses** | **Unvaccinated** | **Two doses** | **Three doses** |
| **Number of individuals included** | 652,301 | 409,082 | 961,899 | 1,725,943 | 65,980 | 22,095 | 545,788 | 84,793 | 179,566 | 1,189,527 | 983,635 | 10,897 | 48,005 | 471,733 |
| **Sex, n (%)** |  |  |  |  |  |  |  |  |  |  |  |  |  |  |
| Men | 310,35<br>(47.6) | 185,597<br>(45.4) | 466,534<br>(48.5) | 816,101<br>(47.3) | 17,255<br>(26.2) | 9,519<br>(43.1) | 256,503<br>(47.0) | 36,104<br>(42.6) | 88,749<br>(49.4) | 580,565<br>(48.8) | 461,281<br>(46.9) | 4,377<br>(40.2) | 23,399<br>(48.7) | 222,216<br>(47.1) |
| Women | 341,942<br>(52.4) | 223,485<br>(54.6) | 495,365<br>(51.5) | 909,842<br>(52.7) | 48,725<br>(73.8) | 12,576<br>(56.9) | 289,285<br>(53.0) | 48,689<br>(57.4) | 90,817<br>(50.6) | 608,962<br>(51.2) | 522,354<br>(53.1) | 6,520<br>(59.8) | 24,606<br>(51.3) | 249,517<br>(52.9) |
| **Median age (IQR)** | 68<br>(64; 74) | 73<br>(69; 77) | 29<br>(21; 35) | 37<br>(24; 48) | 47<br>(38; 54) | 67<br>(62; 74) | 69<br>(64; 75) | 75<br>(66; 84) | 29<br>(21; 39) | 29<br>(19; 39) | 44<br>(32; 52) | 66<br>(62; 74) | 64<br>(61; 69) | 69<br>(64; 76) |
| **Geographical region, n (%)** |  |  |  |  |  |  |  |  |  |  |  |  |  |  |
| Capital Region of Denmark | 176,524<br>(27.1) | 131,694<br>(32.2) | 311,441<br>(32.4) | 586,565<br>(34.0) | 22,191<br>(33.6) | 7,302<br>(33.0) | 161,524<br>(29.6) | 29,034<br>(34.2) | 60,636<br>(33.8) | 365,577<br>(30.7) | 312,193<br>(31.7) | 3,469<br>(31.8) | 10,994<br>(22.9) | 132,175<br>(28.0) |
| Central Denmark Region | 146,827<br>(22.5) | 87,301<br>(21.3) | 227,484<br>(23.6) | 390,709<br>(22.6) | 14,208<br>(21.5) | 3,967<br>(18.0) | 109,593<br>(20.1) | 16,738<br>(19.7) | 36,201<br>(20.2) | 280,700<br>(23.6) | 235,277<br>(23.9) | 2,076<br>(19.1) | 8,946<br>(18.6) | 105,104<br>(22.3) |
| Northern Denmark Region | 75,954<br>(11.6) | 41,881<br>(10.2) | 88,096<br>(9.2) | 164,329<br>(9.5) | 6,366<br>(9.6) | 2,036<br>(9.2) | 61,024<br>(11.2) | 8,163<br>(9.6) | 16,130<br>(9.0) | 124,310<br>(10.5) | 94,906<br>(9.6) | 1,011<br>(9.3) | 7,030<br>(14.6) | 49,839<br>(10.6) |
| Region Zealand | 98,855<br>(15.2) | 56,828<br>(13.9) | 118,070<br>(12.3) | 228,836<br>(13.3) | 8,886<br>(13.5) | 3,728<br>(16.9) | 90,175<br>(16.5) | 13,131<br>(15.5) | 28,034<br>(15.6) | 157,087<br>(13.2) | 132,258<br>(13.4) | 1,911<br>(17.5) | 9,457<br>(19.7) | 73,576<br>(15.6) |
| Region of Southern Denmark | 154,141<br>(23.6) | 91,378<br>(22.3) | 216,808<br>(22.5) | 355,504<br>(20.6) | 14,329<br>(21.7) | 5,062<br>(22.9) | 123,472<br>(22.6) | 17,727<br>(20.9) | 38,565<br>(21.5) | 261,853<br>(22.0) | 209,001<br>(21.2) | 2,430<br>(22.3) | 11,578<br>(24.1) | 111,039<br>(23.5) |
| **Vaccine product, n (%)** |  |  |  |  |  |  |  |  |  |  |  |  |  |  |
| BNT162b2 mRNA (Pfizer/BioNTech) | 0<br>(0.0) | 378,474<br>(92.5) | 0<br>(0.0) | 1,464,639<br>(84.9) | 64,691<br>(98.0) | 0<br>(0.0) | 499,477<br>(91.5) | 80,527<br>(95.0) | 0<br>(0.0) | 970,777<br>(81.6) | 858,689<br>(87.3) | 0<br>(0.0) | 42,829<br>(89.2) | 433,412<br>(91.9) |
| mRNA-1273 (Moderna) | 0<br>(0.0) | 30,608<br>(7.5) | 0<br>(0.0) | 261,304<br>(15.1) | 1,289 (2.0) | 0<br>(0.0) | 46,311<br>(8.5) | 4,266<br>(5.0) | 0<br>(0.0) | 218,750<br>(18.4) | 124,946<br>(12.7) | 0<br>(0.0) | 5,176<br>(10.8) | 38,321<br>(8.1) |

Individuals were able to contribute follow-up time in more than one time category.
\*18-59 years for three doses.

From a high proportion of all SARS-CoV-2 infections, with a median of 77% and 87% mid (<3,000 Ns) and high quality (>150 Ns) genomes were obtained using whole-genome sequencing (WGS) during the Alpha and Delta periods (Fig 1) as previously described (22). A lower median proportion was whole-genome sequenced during the Omicron period (6%) (Fig 1) due to the very high SARS-CoV-2 infection rate and a WGS capacity of 15,000 samples per week (23). However, samples were randomly selected for WGS by an algorithm from all positive samples with cycle threshold (Ct) values below 35.

The elderly were the first to be vaccinated with both the second and the third dose (Fig 2). Overall, the vaccination coverage was high in the study population. The vaccination coverage on January 31, 2022 for two doses was 85% among individuals aged 12-59 and 95% in those 60 years or older. The vaccination coverage on January 31, 2022 for three doses was 64% and 90% among individuals aged 18-59 and 60 years or above, respectively. Therefore, few individuals contributed with follow-up time as unvaccinated (reference group), especially in the Delta and Omicron dominant periods (Fig 2).

**Fig 2.**
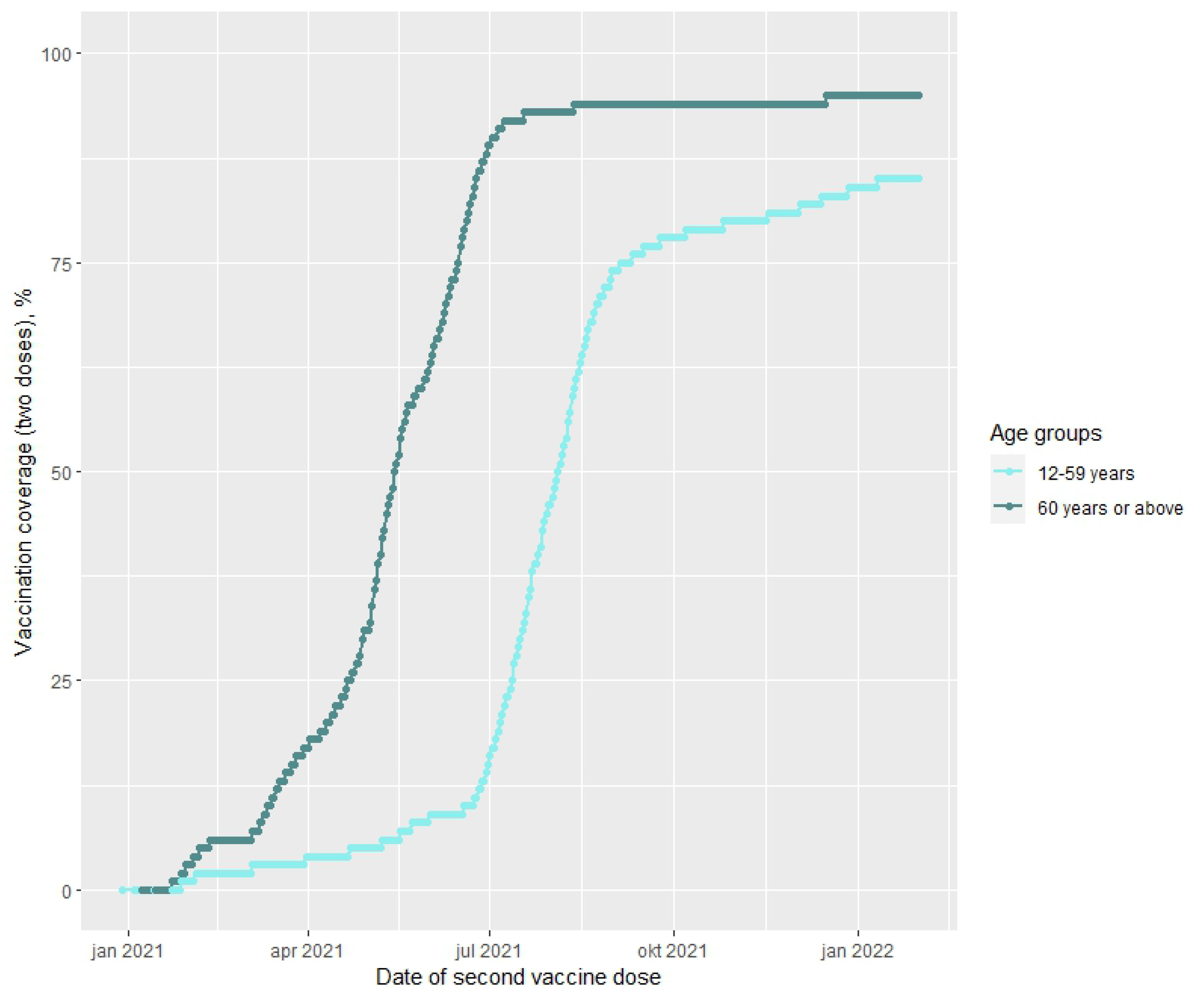
Percentage vaccinated with two doses of BNT162b2 mRNA or mRNA-1273 by age groups.

**Fig 3.**
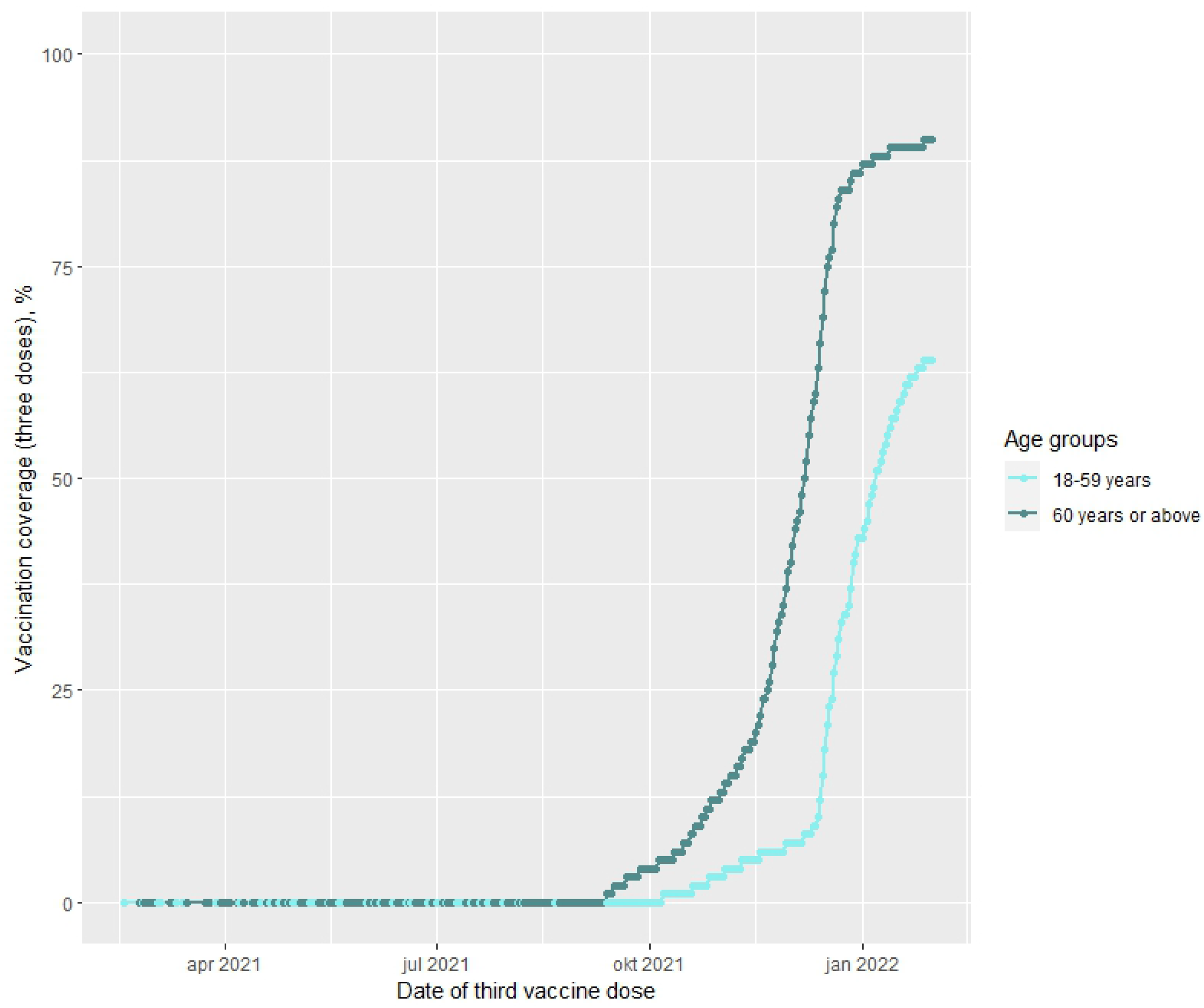
Percentage vaccinated with three doses of BNT162b2 mRNA or mRNA-1273 by age groups.

### Vaccine effectiveness against SARS-CoV-2 infection after two doses

In the Alpha dominant period among individuals aged 60 or above, VE after two doses was 91.0% (95% CI: 88.5; 92.9) 14-30 days since vaccination. The estimates for the subsequent time periods decreased and were 71.5% (95% CI: 54.7; 82.1) >120 days since vaccination. In comparison, the two dose VE against SARS-CoV-2 infection with the Delta variant in individuals aged 60 years or above was 82.2% (95% CI: 75.3; 87.1) at 14-30 days since vaccination. Subsequently, the VE estimates decreased to 49.8% (95% CI: 46.5; 52.8) >120 days since vaccination. Slightly higher estimates were observed among individuals aged 12-59 years, where VE against SARS-CoV-2 infection with the Delta variant was 92.2% (95% CI: 91.8; 92.6) 14-30 days since vaccination and decreased to 64.9% (95% CI: 64.0; 65.8) >120 days since vaccination. Markedly lower two dose VE estimates were observed against SARS-CoV-2 infection with the Omicron variant for both age groups. Among individuals aged 12-59 years, VE was 39.8% (95% CI: 38.4; 41.2) 14-30 days after vaccination. The estimates decreased to 13.2% (95% CI: 12.5; 13.9) >120 days since vaccination. Similar estimates but with wider CIs were observed among individuals aged 60 years or above (Table 2 and Fig 4).

**Table 2.** Adjusted vaccine effectiveness of two doses BNT162b2 mRNA or mRNA-1273 against SARS-CoV-2 infection with the Alpha, Delta and Omicron variants by age groups (12-59 years and 60 years or above).

|  | Alpha |  |  |  |  | Delta |  |  |  |  | Omicron |  |  |  |  |
| --- | --- | --- | --- | --- | --- | --- | --- | --- | --- | --- | --- | --- | --- | --- | --- |
|  | Population | Person-years | Cases | Adjusted VE | 95% CI | Population | Person-years | Cases | Adjusted VE | 95% CI | Population | Person-years | Cases | Adjusted VE | 95% CI |
| **12-59 years** |  |  |  |  |  |  |  |  |  |  |  |  |  |  |  |
| Unvaccinated |  |  |  |  |  | 961,900 | 143,391 | 43,583 | 1<br>(reference) |  | 179,566 | 15,483 | 96,239 | 1<br>(reference) |  |
| Time since vaccination |  |  |  |  |  |  |  |  |  |  |  |  |  |  |  |
| 14-30 days |  |  |  |  |  | 1,600,368 | 74,050 | 1,623 | 92.2 | 91.8; 92.6 | 61,458 | 2,147 | 8,245 | 39.8 | 38.4; 41.2 |
| 31-60 days |  |  |  |  |  | 1,598,448 | 129,640 | 3,718 | 88.1 | 87.7; 88.5 | 63,908 | 3,018 | 16,503 | 31.6 | 30.5; 32.8 |
| 61-90 days |  |  |  |  |  | 1,581,102 | 123,576 | 7,873 | 80.8 | 80.3; 81.3 | 57,574 | 2,157 | 9,759 | 32.1 | 30.6; 33.5 |
| 91-120 days |  |  |  |  |  | 1,400,933 | 85,889 | 14,950 | 72.2 | 71.5; 72.8 | 221,118 | 7,015 | 22,907 | 31.4 | 30.3; 32.4 |
| >120 days |  |  |  |  |  | 750,408 | 59,185 | 10,359 | 64.9 | 64.0; 65.8 | 1,075,946 | 56,886 | 276,028 | 13.2 | 12.5; 13.9 |
| **60 years or above** |  |  |  |  |  |  |  |  |  |  |  |  |  |  |  |
| Unvaccinated | 652,301 | 111,187 | 4462 | 1<br>(reference) |  | 22,095 | 6,894 | 1,113 | 1<br>(reference) |  | 10,897 | 1,051 | 3,354 | 1<br>(reference) |  |
| Time since vaccination |  |  |  |  |  |  |  |  |  |  |  |  |  |  |  |
| 14-30 days | 407,491 | 16,796 | 78 | 91 | 88.5; 92.9 | 199,219 | 6,996 | 59 | 82.2 | 75.3; 87.1 | 1,340 | 48 | 96 | 39.9 | 26.4; 50.9 |
| 31-60 days | 323,574 | 16,189 | 145 | 83.1 | 79.6; 86.0 | 360,046 | 21,964 | 393 | 74.3 | 69.9; 78.0 | 1,289 | 61 | 136 | 39.2 | 27.8; 48.8 |
| 61-90 days | 116,297 | 7,274 | 119 | 72.6 | 66.3; 77.8 | 447,280 | 34,207 | 687 | 77.4 | 74.4; 80.0 | 1,071 | 43 | 102 | 26.4 | 10.4; 39.6 |
| 91-120 days | 58,340 | 3,467 | 49 | 82.7 | 76.5; 87.2 | 496,174 | 38,464 | 1,161 | 69.8 | 66.7; 72.6 | 1,750 | 77 | 173 | 24.4 | 11.9; 35.1 |
| >120 days | 35,698 | 1,445 | 24 | 71.5 | 54.7; 82.1 | 534,299 | 92,240 | 12,198 | 49.8 | 46.5; 52.8 | 45,844 | 1,783 | 4,722 | 4.7 | 0.2; 8.9 |

VE = vaccine effectiveness. CI = confidence intervals. Person-years in days. VE estimates adjusted for five-year age groups, sex, and geographical region. Individuals were able to contribute follow-up time in more than one time category and (if vaccinated during the study period) to both the analysis of VE after two and three doses.

**Fig 4.**
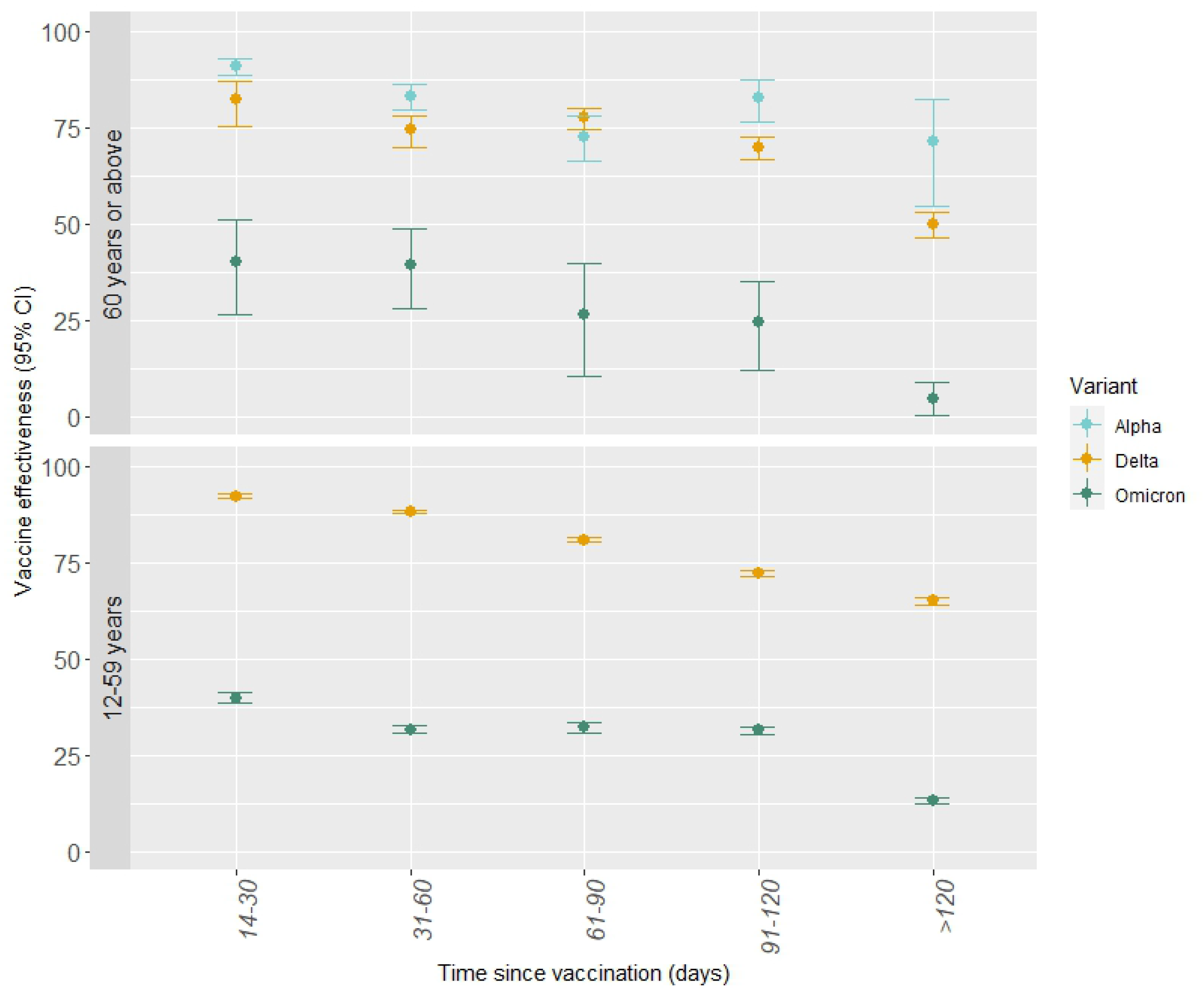
Adjusted vaccine effectiveness against SARS-CoV-2 infection after two doses BNT162b2 mRNA or mRNA-1273 by SARS-CoV-2 variants and age groups.

### Vaccine effectiveness against SARS-CoV-2 infection after three doses

Similar to the VE estimates after two doses, the VE after three doses was markedly lower against SARS-CoV-2 infection with the Omicron variant than that against the Alpha and Delta variants, although the VE against the Omicron variant was higher after three than after two doses and with less waning by time since vaccination (Table 2 and Table 3). Among individuals aged 18-59 years, VE against SARS-CoV-2 infection with the Delta variant was 89.5% (95% CI: 87.7; 91.0) 14-30 days since vaccination and 83.5% (95% CI: 69.4; 91.1) 61-90 days since vaccination. Similar estimates were observed among individuals aged 60 years or above. In comparison, for individuals aged 18-59 years, VE after three doses against SARS-CoV-2 infection with the Omicron variant was 55.2% (95% CI: 54.7; 55.6) 14-30 days since vaccination and 49.9% (95% CI: 46.5; 53.1) >120 days since vaccination. Among individuals aged 60 years or above, VE against SARS-CoV-2 infection with the Omicron variant after three doses was 57.6% (95% CI: 55.8; 59.4) 14-30 days since vaccination and 52.8% (95% CI: 49.3; 56.0) >120 days since vaccination (Table 3 and Fig 5).

**Table 3.** Adjusted vaccine effectiveness of three doses BNT162b2 mRNA or mRNA-1273 against SARS-CoV-2 infection with the Delta and Omicron variants by age groups (18-59 years and 60 years or above).

|  | Delta |  |  |  |  | Omicron |  |  |  |  |
| --- | --- | --- | --- | --- | --- | --- | --- | --- | --- | --- |
|  | Population | Person-years | Cases | Adjusted VE | 95% CI | Population | Person-years | Cases | Adjusted VE | 95% CI |
| **18-59 years** |  |  |  |  |  |  |  |  |  |  |
| Unvaccinated | 757,824 | 110,495 | 34,414 | 1 (reference) |  | 145,080 | 12,664 | 74,692 | 1 (reference) |  |
| Time since vaccination |  |  |  |  |  |  |  |  |  |  |
| 14-30 days | 65,955 | 2,200 | 155 | 89.5 | 87.7; 91.0 | 892,781 | 34,645 | 105,187 | 55.2 | 54.7; 55.6 |
| 31-60 days | 22,694 | 718 | 93 | 77.6 | 72.4; 81.7 | 641,642 | 20,388 | 89,239 | 50.8 | 50.2; 51.4 |
| 61-90 days | 4,087 | 74 | 10 | 83.5 | 69.4; 91.1 | 93,683 | 5,590 | 15,754 | 52.9 | 52.0; 53.7 |
| 91-120 days |  |  |  |  |  | 47,530 | 1,537 | 5,902 | 51.1 | 49.7; 52.5 |
| >120 days |  |  |  |  |  | 6,415 | 227 | 979 | 49.9 | 46.5; 53.1 |
| **60 years or above** |  |  |  |  |  |  |  |  |  |  |
| Unvaccinated | 22,095 | 6,894 | 1,113 | 1 (reference) |  | 10,897 | 1,051 | 3,354 | 1 (reference) |  |
| Time since vaccination |  |  |  |  |  |  |  |  |  |  |
| 14-30 days | 84,768 | 3,183 | 161 | 86 | 83.3; 88.3 | 335,455 | 12,712 | 13,485 | 57.6 | 55.8; 59.4 |
| 31-60 days | 48,428 | 2,612 | 188 | 82.9 | 79.5; 85.7 | 390,857 | 21,089 | 34,164 | 55.3 | 53.6; 56.9 |
| 61-90 days | 15,844 | 258 | 34 | 81.2 | 72.9; 87.0 | 177,842 | 7,035 | 10,409 | 58.3 | 56.5; 60.0 |
| 91-120 days |  |  |  |  |  | 78,411 | 3,947 | 5,361 | 56.6 | 54.4; 58.7 |
| >120 days |  |  |  |  |  | 34,293 | 1,215 | 2,428 | 52.8 | 49.3; 56.0 |

VE = vaccine effectiveness. CI = confidence intervals. VE estimates adjusted for five-year age groups, sex, and geographical region. Individuals were able to contribute follow-up time in more than one time category and (if vaccinated during the study period) to both the analysis of VE after two and three doses.

**Fig 5.**
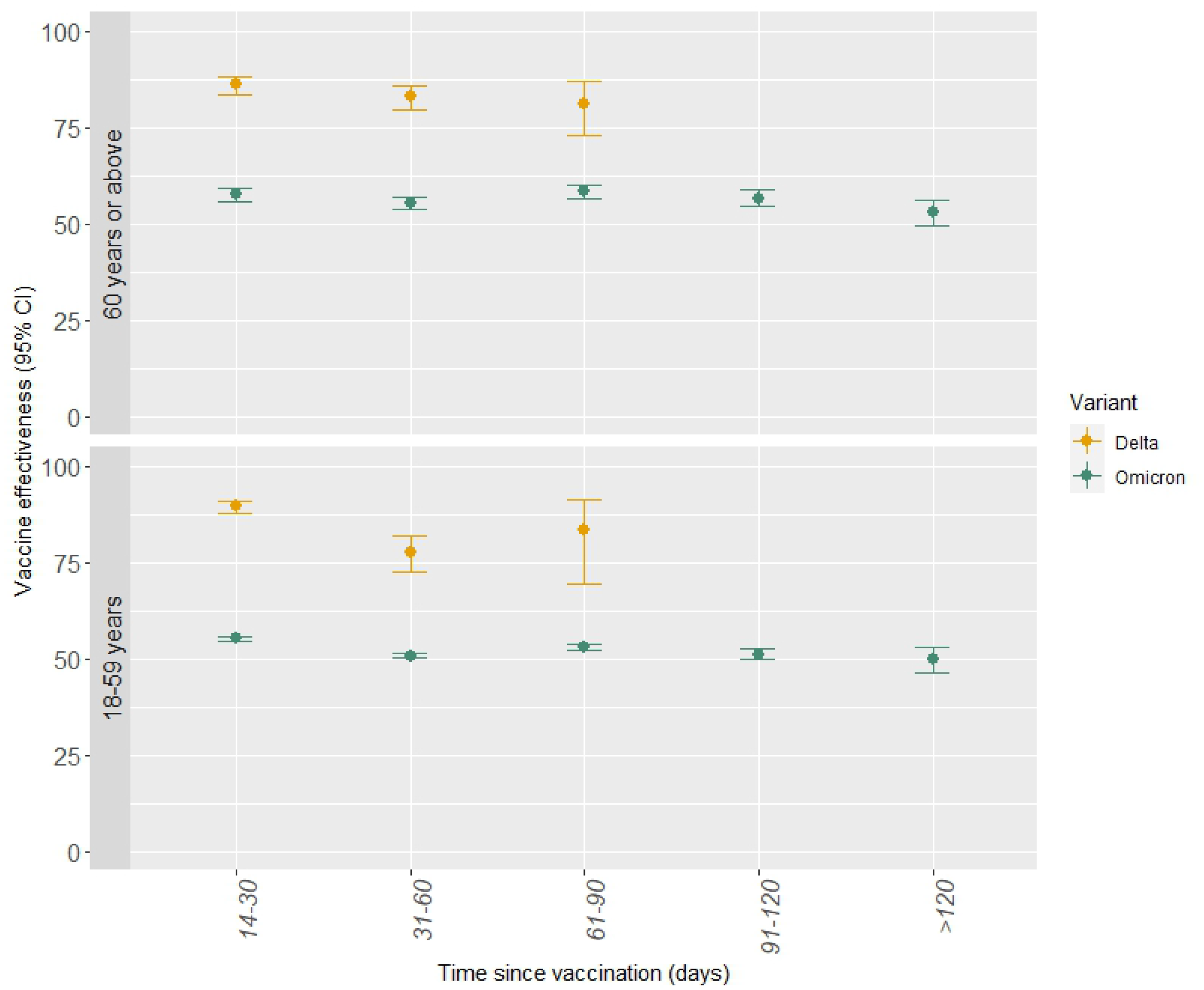
Adjusted vaccine effectiveness against SARS-CoV-2 infection after three doses BNT162b2 mRNA or mRNA-1273 by SARS-CoV-2 variants and age groups.

### Vaccine effectiveness against COVID-19-related hospitalization after two doses

A high VE against COVID-19-related hospitalization after two doses was observed for both the Alpha and Delta variants. However, only limited protection was observed against COVID-19-related hospitalization following infection with the Omicron variant (Table 4). Among individuals aged 60 years or above, VE against COVID-19-related hospitalization with the Alpha and Delta variants after two doses was 96.4% (95% CI: 92.6; 98.3) and 100% (95% CI was not estimated as no hospital admissions were observed), respectively, 14-30 days since vaccination. The estimates decreased to 90.5% (Alpha, 95% CI: 67.0; 97.2) and 86.2% (Delta, 95% CI: 84.2; 87.9) >120 days since vaccination. For the Delta variant, similar estimates were observed among individuals aged 12-59 years (Table 4). Among individuals aged 12-59 years, VE against COVID-19-related hospitalization following infection with the Omicron variant was 62.4% (95% CI: 46.3; 73.6) 14-30 days since vaccination and 65.9% (95% CI: 62.0; 69.4) >120 days since vaccination. It was not possible to estimate two dose VE against COVID-19-related hospitalization following infection with the Omicron variant among individuals aged 60 years or above due to few cases, and since the majority of this group had already received a third vaccine dose at this time (Table 4 and Fig 6).

**Table 4.** Adjusted vaccine effectiveness of two doses BNT162b2 mRNA or mRNA-1273 against COVID-19-related hospitalization following infection with the Alpha, Delta and Omicron variants by age groups (12-59 years and 60 years or above)

|  | Alpha |  |  |  |  | Delta |  |  |  |  | Omicron |  |  |  |  |
| --- | --- | --- | --- | --- | --- | --- | --- | --- | --- | --- | --- | --- | --- | --- | --- |
|  | Population | Person-years | Cases | Adjusted VE | 95% CI | Population | Person-years | Cases | Adjusted VE | 95% CI | Population | Person-years | Cases | Adjusted VE | 95% CI |
| **12-59 years** |  |  |  |  |  |  |  |  |  |  |  |  |  |  |  |
| Unvaccinated |  |  |  |  |  | 961,900 | 143,391 | 934 | 1<br>(reference) |  | 179,566 | 15,483 | 725 | 1<br>(reference) |  |
| Time since vaccination |  |  |  |  |  |  |  |  |  |  |  |  |  |  |  |
| 14-30 days |  |  |  |  |  | 1,600,368 | 74,050 | 6 | 99.1 | 98.0; 99.6 | 61,458 | 2,147 | 32 | 62.4 | 46.3; 73.6 |
| 31-60 days |  |  |  |  |  | 1,598,448 | 129,640 | 21 | 98.3 | 97.4; 98.9 | 63,908 | 3,018 | 67 | 51.4 | 37.4; 62.3 |
| 61-90 days |  |  |  |  |  | 1,581,102 | 123,576 | 18 | 98.4 | 97.4; 99.0 | 57,574 | 2,157 | 36 | 59.8 | 43.7; 71.3 |
| 91-120 days |  |  |  |  |  | 1,400,933 | 85,889 | 58 | 95.9 | 94.6; 96.9 | 221,118 | 7,015 | 84 | 61.4 | 51.3; 69.4 |
| >120 days |  |  |  |  |  | 750,408 | 59,185 | 115 | 91.6 | 89.5; 93.2 | 1,075,946 | 56,886 | 673 | 65.9 | 62.0; 69.4 |
| **60 years or above** |  |  |  |  |  |  |  |  |  |  |  |  |  |  |  |
| Unvaccinated | 652,301 | 111,187 | 658 | 1<br>(reference) |  | 22,095 | 6,894 | 287 | 1<br>(reference) |  |  |  |  |  |  |
| Time since vaccination |  |  |  |  |  |  |  |  |  |  |  |  |  |  |  |
| 14-30 days | 407,491 | 16,796 | 8 | 96.4 | 92.6; 98.3 | 199,219 | 6,996 | 0 | 100 | * |  |  |  |  |  |
| 31-60 days | 323,574 | 16,189 | 22 | 92.1 | 87.4; 95.1 | 360,046 | 21,964 | 8 | 97.7 | 95.2; 98.9 |  |  |  |  |  |
| 61-90 days | 116,297 | 7,274 | 25 | 82.5 | 72.1; 89.0 | 447,280 | 34,207 | 24 | 97.1 | 95.5; 98.1 |  |  |  |  |  |
| 91-120 days | 58,340 | 3,467 | 8 | 91 | 80.9; 95.7 | 496,174 | 38,464 | 44 | 96.3 | 94.7; 97.3 |  |  |  |  |  |
| >120 days | 35,698 | 1,445 | 3 | 90.5 | 67.0; 97.2 | 534,299 | 92,240 | 855 | 86.2 | 84.2; 87.9 |  |  |  |  |  |

VE = vaccine effectiveness. CI = confidence intervals. VE estimates adjusted for five-year age groups, sex, and geographical region. Individuals were able to contribute follow-up time in more than one time category and (if vaccinated during the study period) to both the analysis of VE after two and three doses.
\* It was not possible within the model to estimate a 95% confidence interval for the estimated vaccine effectiveness against COVID-19-related hospitalization with the Delta variant 14-30 days after the second dose as no COVID-19-related hospitalization were observed.

**Fig 6.**
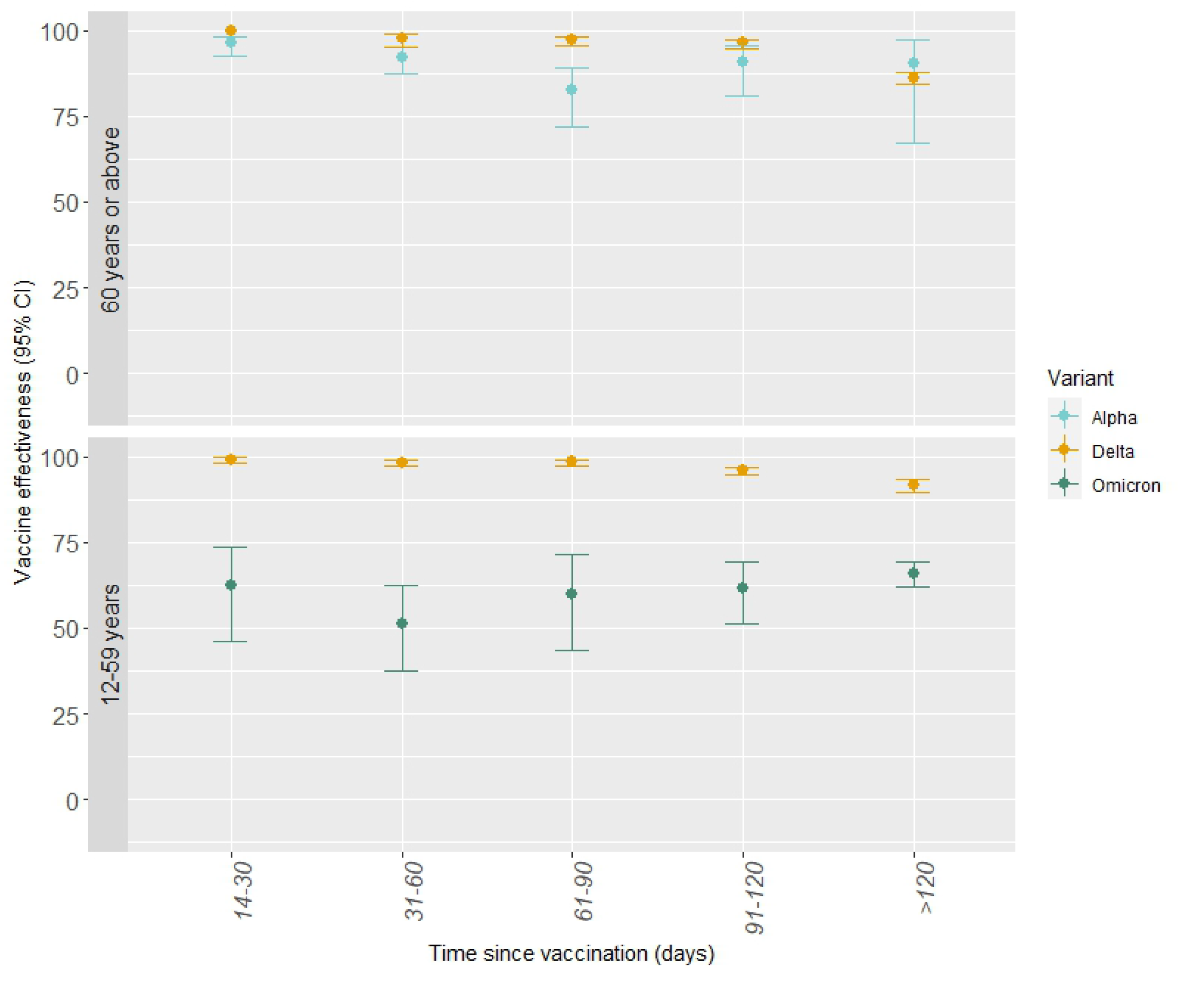
Adjusted vaccine effectiveness against COVID-19-related hospitalization after two doses BNT162b2 mRNA or mRNA-1273 by SARS-CoV-2 variants and age groups.

### Vaccine effectiveness against COVID-19-related hospitalization after three doses

For both age groups, small absolute differences were observed in the VE estimates of three doses against COVID-19-related hospitalization between the Delta and Omicron variants. However, no waning was observed for the Delta variant while the estimates indicated waning over time against COVID-19-related hospitalization following infection with the Omicron variant. However, the available time interval after vaccination during the Delta period was shorter. In individuals aged 18-59 years, VE against COVID-19-related hospitalization following infection with the Delta variant was 94.8% (95% CI: 85.9; 98.1) 14-30 days and 68.4% (95% CI: 41.4; 83.0) 31-60 days since vaccination. In individuals aged 60 years or above, VE against COVID-19-related hospitalization following infection with the Delta variant was 96.6% (95% CI: 93.9; 98.1) 14-30 days and 91.4% (95% CI: 79.8; 96.4) 61-90 days since vaccination. In comparison, in individuals aged 18-59 years, VE against COVID-19-related hospitalization following infection with the Omicron variant was 89.8% (95% CI: 87.9; 91.3) 14-30 days since vaccination. From then on, a gradual decline in VE was observed reaching 33.3% (95% CI: 0.9; 55.1) >120 days since vaccination. A smaller decrease in the VE against COVID-19-related hospitalization following infection with the Omicron was observed among individuals aged 60 years or above with an estimated VE of 94.4% (95% CI: 93.0; 95.5) 14-30 days and 77.3% (95% CI: 70.9; 82.3) >120 days since vaccination (Table 5 and Fig 7).

**Table 5.** Adjusted vaccine effectiveness of three doses BNT162b2 mRNA or mRNA-1273 against COVID-19-related hospitalization following infection with the Delta and Omicron variants by age groups (18-59 years and ≥60 years)

|  | Delta |  |  |  |  | Omicron |  |  |  |  |
| --- | --- | --- | --- | --- | --- | --- | --- | --- | --- | --- |
|  | Population | Person-years | Cases | Adjusted VE | 95% CI | Population | Person-years | Cases | Adjusted VE | 95% CI |
| **18-59 years** |  |  |  |  |  |  |  |  |  |  |
| Unvaccinated | 757,824 | 110,495 | 911 | 1 (reference) |  | 145,080 | 12,664 | 681 | 1 (reference) |  |
| Time since vaccination |  |  |  |  |  |  |  |  |  |  |
| 14-30 days | 65,955 | 2,200 | 4 | 94.8 | 85.9; 98.1 | 892,781 | 34,645 | 210 | 89.8 | 87.9; 91.3 |
| 31-60 days | 22,694 | 718 | 11 | 68.4 | 41.4; 83.0 | 641,642 | 20,388 | 216 | 87.4 | 84.9; 89.6 |
| 61-90 days |  |  |  |  |  | 93,683 | 5,590 | 97 | 80.5 | 75.4; 84.5 |
| 91-120 days |  |  |  |  |  | 47,530 | 1,537 | 61 | 72.8 | 63.6; 79.7 |
| >120 days |  |  |  |  |  | 6,415 | 227 | 34 | 33.3 | 0.9; 55.1 |
| **60 years or above** |  |  |  |  |  |  |  |  |  |  |
| Unvaccinated | 22,095 | 6,894 | 287 | 1 (reference) |  | 10,897 | 1,051 | 298 | 1 (reference) |  |
| Time since vaccination |  |  |  |  |  |  |  |  |  |  |
| 14-30 days |  |  |  |  |  | 335,455 | 12,712 | 136 | 94.4 | 93.0; 95.5 |
| 31-60 days | 84,768 | 3,183 | 13 | 96.6 | 93.9; 98.1 | 390,857 | 21,089 | 366 | 93.8 | 92.7; 94.7 |
| 61-90 days | 48,428 | 2,612 | 44 | 89.1 | 84.3; 92.4 | 177,842 | 7,035 | 272 | 91.7 | 90.2; 93.0 |
| 91-120 days | 15,844 | 258 | 6 | 91.4 | 79.8; 96.4 | 78,411 | 3,947 | 353 | 84.5 | 81.6; 87.0 |
| >120 days |  |  |  |  |  | 34,293 | 1,215 | 176 | 77.3 | 70.9; 82.3 |

VE = vaccine effectiveness. CI = confidence intervals. VE estimates adjusted for five-year age groups, sex, and geographical region. Individuals were able to contribute follow-up time in more than one time category and (if vaccinated during the study period) to both the analysis of VE after two and three doses.

**Fig 7.**
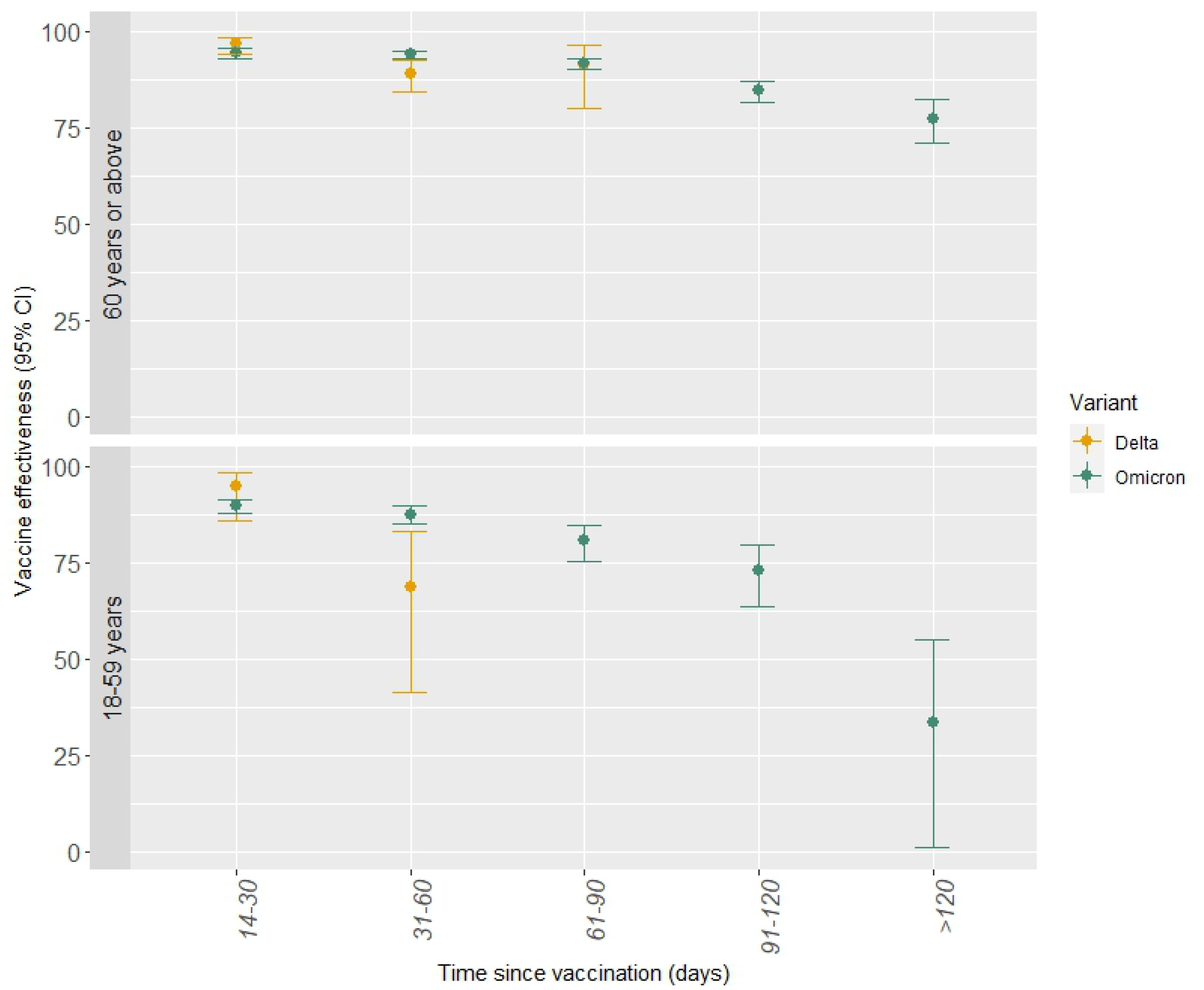
Adjusted vaccine effectiveness against COVID-19-related hospitalization after three doses BNT162b2 mRNA or mRNA-1273 by SARS-CoV-2 variants and age groups.

## Discussion

Compared to unvaccinated individuals, vaccination with two or three doses of BNT162b2 mRNA or mRNA-1273 was associated with protection against infection with the Alpha, Delta and Omicron SARS-CoV-2 variants, peaking 14-30 days since vaccination with either two or three doses. However, the protection against infection with the Omicron variant was markedly lower compared to infection with the Alpha or Delta variants. The protection against infection with the three variants afforded by vaccination after two doses decreased with time since vaccination. However, the VE against infection with the Delta and Omicron variants decreased less over time after three doses. High protection and less pronounced waning were observed against COVID-19-related hospitalization following infection with the Alpha and Delta variants after two doses. However, the third dose contributed to greater levels of protection against COVID-19-related hospitalization following infection especially with the Omicron variant. It was unexpected that individuals aged 12-59 years had slightly lower VE against infection with the Omicron variant in all time intervals compared to individuals aged 60 years or above. This may be explained by biases introduced in non-randomized studies including differences in behavior between unvaccinated and vaccinated individuals (24).

Our results for the VE against SARS-CoV-2 infection with the Alpha and Delta variants align with those of a test-negative case-control study from England that observed a slightly higher VE against SARS-CoV-2 infection with the Alpha variant compared to the Delta variant after two doses of the BNT162b2 mRNA vaccine (4). They observed a VE against SARS-CoV-2 infection with the Alpha and Delta variants of 93.7% (95% CI: 91.6; 95.3) and 88.0% (95% CI: 85.3; 90.1), respectively (4). However, in a study from the UK, similar VE estimates against infection with the Alpha (78% (95% CI: 68; 84)) or Delta (80% (95% CI: 77; 83)) variants after two doses of BNT162b2 mRNA were observed (5). Previous studies have observed markedly higher protection against infection with the Delta variant compared to the Omicron variant (6-8). A test-negative case-control study from Southern California found that VE against infection with the Delta variant after two doses of mRNA-1273 was high and waned slowly with VE of 80.2% (95% CI: 68.2; 87.7) and 61.3% (95% CI: 55.0; 66.7) 14-90 days and >270 days since vaccination, respectively (8). Furthermore, they observed VE against hospitalization with the Delta variant of ≥99% after two and three doses (8). Similar to our study, a test-negative case-control study from England also observed waning over time and that a third dose contributed to greater levels of protection (7). They observed that VE against infection with the Delta variant after two doses BNT162b2 mRNA was 90.9% (95% CI: 89.6; 92.0) 2-4 weeks since vaccination and declined to 62.7% (95% CI: 61.6; 63.7) ≥25 weeks since vaccination. After the third dose, the VE against infection with the Delta variant was 95.1% (95% CI: 94.8; 95.4) 2-4 weeks since vaccination and declined to 89.9% (95% CI: 89.2; 90.5) ≥10 weeks since vaccination (7). The VE was higher with the mRNA-1273 vaccine (7).

For the Omicron variant, the study from Southern California observed only a modest protection of 44.0% (95% CI: 35.1; 51.6) against infection 14-90 days since vaccination with two doses mRNA-1273 (8). The VE decreased quickly thereafter to 23.5% (95% CI: 16.4; 30.0) 91-180 days and 5.9% (95% CI: 0.04-11.0) >270 days since vaccination (8). Similar to our results, the VE against infection with the Omicron variant increased after the third dose (8). However, they observed higher VE estimates against hospitalization with the Omicron variant compared to our study with estimates of 84.5% (95% CI: 23.0; 96.9) and 99.2% (95% CI: 76.3; 100.0) after two and three doses, respectively (8).

A test-negative case-control study from England observed limited protection against symptomatic disease caused by the Omicron variant but reported that a third dose substantially increased protection (7). Their observed VE estimates were higher and waned faster after the third dose than observed in our study. VE against infection with the Omicron variant was 65.5% (95% CI: 63.9; 67.0) and 8.8% (95% CI: 7.0; 10.5) 2-4 weeks and ≥25 weeks since vaccination, respectively (7). After the third dose, VE against infection with the Omicron variant was 67.2% (95% CI: 66.5; 67.8) and 45.7% (95% CI: 44.7; 46.7) 2-4 weeks and ≥10 weeks since vaccination, respectively (7). An explanation to the lower estimates in our study may be that we estimated VE against SARS-CoV-2 infection regardless of the symptom status.

## Strengths and limitations

The strengths of this study are the large scale of testing for SARS-CoV-2 including unlimited and easily accessible free-of-charge RT-PCR tests, as well as the ability to individually link data on all residents in Denmark across the nationwide high-quality registries. The high sensitivity (97.1%) and specificity (99.98%) observed for the RT-PCR test (21) minimize the risk of misclassification of the outcome. However, we were not able to discriminate between asymptomatic and symptomatic infections. In addition, a previous study has observed an inherent increased transmissibility of the Omicron sub-linage BA.2 (22) and we cannot rule out that the VE differs between the Omicron sub-linages. BA.1 and BA.2 was the most frequent Omicron sub-linages in Denmark (25). BA.1 was most prevalent in the beginning of the included Omicron dominated period. However, the prevalence of BA.2 has been increasing faster than BA.1 (25). Due to the short BA.1 dominated period, it was not possible to separate the VE analysis by BA.1 and BA.2.

COVID-19-related hospitalization was defined as an all-cause hospital admission lasting at least 12 hours and occurring up to 14 days after or two days before a positive SARS-CoV-2 test. Some of the hospitalizations included in our analysis will therefore have been caused by other factors than COVID-19. Assuming that the rate of hospitalization due to other causes is similar in the vaccinated and unvaccinated populations, the lack of specificity in the definition may have resulted in an underestimation of the VE. In addition, an effort was made to ensure equal access to COVID-19 vaccination for all Danish residents. This was done through an online booking system, special campaigns, offering vaccination in some workplaces, translating the information about COVID-19 vaccination to several languages and arranging transport and pop-up vaccination for those who were not able to reach the vaccination clinics on their own. However, the populations initially prioritized for COVID-19 vaccination were the most vulnerable citizens and frontline healthcare workers whereas the younger population was invited later. Therefore, it was not possible to estimate VE of two and three doses for both age groups in all defined periods. No individuals had received the third dose in the Alpha dominant period and the majority of the oldest age group had already received third vaccine dose in the Omicron dominant period.

In general, non-randomized studies assessing COVID-19 VE can easily be flawed (24), which may also apply to this study. Although we adjusted the Cox regression models for potential confounders (calendar time, age, sex and geographical region), we cannot exclude time-varying factors such as test frequency and differences in behavior or adherence to COVID-19 guidelines between vaccinated and unvaccinated, which may impact infection exposure. Vaccinated individuals may be more frequently tested (and thus more likely to document infection) if they are more health-conscious compared with unvaccinated individuals. The vaccinated individuals may be less frequently tested, if the vaccination reduces the severity of the infection and thus fewer infected people have symptoms. Furthermore, public health authorities may encourage more frequent testing for the unvaccinated (24). Furthermore, vaccinated individuals may become more heavily exposed to the virus after vaccination, if they feel liberated to engage in activities with more frequent and high-risk exposure (24). This phenomenon of risk compensation decreases the benefit of vaccination (26). However, some data have suggested little change in protective behavior early after vaccination (27).

Overall, this study contributed with evidence of high vaccine protection against SARS-CoV-2 infection and importantly against hospitalization with the Alpha and Delta variants after two doses of BNT162b2 mRNA or mRNA-1273. A third dose seems necessary to maintain protection against infection for longer time and in particular to ensure good protection against COVID-19-related hospitalization with the Omicron variant. In the context of the current Omicron wave, it is notable that a third dose contributed to greater levels of protection against both infection and especially COVID-19-related hospitalization following infection with the Omicron variant.

## Data Availability

Data cannot be shared publicly due data protection regulation. Data are available from the Danish Health Data Authority for researchers who meet the criteria for access to confidential data. The data are available for research upon reasonable request and with permission from the Danish Data Protection Agency and the Danish Health Data Authority: https://sundhedsdatastyrelsen.dk/da/english/health_data_and_registers/research_services

## Funding

Not applicable as neither Statens Serum Institut nor any authors received funding for this study.

## Acknowledgement

The authors are grateful to the Danish Health Data Authority for their help in defining the population. We would also like to thank the Department of Data Integration and Analysis at Statens Serum Institut for data management.

## Notes

### Competing Interest Statement

The authors have declared no competing interest.

### Funding Statement

The author(s) received no specific funding for this work

### Author Declarations

The data becomes or are already available for research upon reasonable request and with permission from the Danish Data Protection Agency and Danish Health Data Authority. Ethics: We used only administrative register data for the study. According to Danish law, ethics approval is exempt for such research, and the Danish Data Protection Agency, which is a dedicated ethics and legal oversight body, thus waives ethical approval for our study of administrative register data, when no individual contact of participants is neccessary and only aggregate results are included as findings. The study is therefore fully compliant with all legal and ethical requirements and there are no further processes available regarding such studies.

## References

1. Barda N, Dagan N, Cohen C, Hernán MA, Lipsitch M, Kohane IS, et al. Effectiveness of a third dose of the BNT162b2 mRNA COVID-19 vaccine for preventing severe outcomes in Israel: an observational study. Lancet (London, England). 2021;398(10316):2093–100.

2. Keehner J, Horton LE, Binkin NJ, Laurent LC, Pride D, Longhurst CA, et al. Resurgence of SARS-CoV-2 Infection in a Highly Vaccinated Health System Workforce. The New England journal of medicine. 2021;385(14):1330–2.

3. Goldberg Y, Mandel M, Bar-On YM, Bodenheimer O, Freedman L, Haas EJ, et al. Waning Immunity after the BNT162b2 Vaccine in Israel. N Engl J Med. 2021;385(24):e85.

4. Lopez Bernal J, Andrews N, Gower C, Gallagher E, Simmons R, Thelwall S, et al. Effectiveness of Covid-19 Vaccines against the B.1.617.2 (Delta) Variant. N Engl J Med. 2021;385(7):585–94.

5. Pouwels KB, Pritchard E, Matthews PC, Stoesser N, Eyre DW, Vihta KD, et al. Effect of Delta variant on viral burden and vaccine effectiveness against new SARS-CoV-2 infections in the UK. Nat Med. 2021;27(12):2127–35.

6. Accorsi EK, Britton A, Fleming-Dutra KE, Smith ZR, Shang N, Derado G, et al. Association Between 3 Doses of mRNA COVID-19 Vaccine and Symptomatic Infection Caused by the SARS-CoV-2 Omicron and Delta Variants. JAMA. 2022;327(7):639–51.

7. Andrews N, Stowe J, Kirsebom F, Toffa S, Rickeard T, Gallagher E, et al. Covid-19 Vaccine Effectiveness against the Omicron (B.1.1.529) Variant. N Engl J Med. 2022.

8. Tseng HF, Ackerson BK, Luo Y, Sy LS, Talarico CA, Tian Y, et al. Effectiveness of mRNA-1273 against SARS-CoV-2 Omicron and Delta variants. Nat Med. 2022.

9. Feikin DR, Higdon MM, Abu-Raddad LJ, Andrews N, Araos R, Goldberg Y, et al. Duration of effectiveness of vaccines against SARS-CoV-2 infection and COVID-19 disease: results of a systematic review and meta-regression. Lancet. 2022.

10. Lauring AS, Tenforde MW, Chappell JD, Gaglani M, Ginde AA, McNeal T, et al. Clinical severity of, and effectiveness of mRNA vaccines against, covid-19 from omicron, delta, and alpha SARS-CoV-2 variants in the United States: prospective observational study. BMJ. 2022;376:e069761.

11. Ritchie H, Mathieu E, Rodés-Guirao L, Appel C, Giattino C, Ortiz-Ospina E, et al. Coronavirus (COVID-19) Cases. Our World in Data.

12. Tracking SARS-CoV-2 variants: World Health Organization; [cited 2022 24 April]. Available from: https://www.who.int/en/activities/tracking-SARS-CoV-2-variants/.

13. Hansen C, Schelde A, Moustsen-Helm I, Emborg H-D, Eriksen R, Stegger M, et al. Vaccine effectiveness against infection and COVID19-associated hospitalisation with the Omicron (B.1.1.529) variant after vaccination with the BNT162b2 or mRNA-1273 vaccine: A nationwide Danish cohort study. Nature Portfolio (Preprint). 2022.

14. Nyberg T, Ferguson NM, Nash SG, Webster HH, Flaxman S, Andrews N, et al. Comparative analysis of the risks of hospitalisation and death associated with SARS-CoV-2 omicron (B.1.1.529) and delta (B.1.617.2) variants in England: a cohort study. Lancet. 2022;399(10332):1303–12.

15. Schmidt M, Pedersen L, Sørensen HT. The Danish Civil Registration System as a tool in epidemiology. Eur J Epidemiol. 2014;29(8):541–9.

16. Target groups for vaccination: The Danish Health Authority; [cited 2021 August 6]. Available from: https://www.sst.dk/en/English/Corona-eng/Vaccination%20against%20COVID-19/Who%20should%20be%20vaccinated.

17. Grove Krause T, Jakobsen S, Haarh M, Mølbak K. The Danish vaccination register. Euro Surveill. 2012;17(17).

18. Schønning K, Dessau RB, Jensen TG, Thorsen NM, Wiuff C, Nielsen L, et al. Electronic reporting of diagnostic laboratory test results from all healthcare sectors is a cornerstone of national preparedness and control of COVID-19 in Denmark. Apmis. 2021;129(7):438–51.

19. Fernandez-Montero A, Argemi J, Rodríguez JA, Ariño AH, Moreno-Galarraga L. Validation of a rapid antigen test as a screening tool for SARS-CoV-2 infection in asymptomatic populations. Sensitivity, specificity and predictive values. EClinicalMedicine. 2021:100954.

20. Schmidt M, Schmidt SA, Sandegaard JL, Ehrenstein V, Pedersen L, Sørensen HT. The Danish National Patient Registry: a review of content, data quality, and research potential. Clin Epidemiol. 2015;7:449–90.

21. von Elm E, Altman DG, Egger M, Pocock SJ, Gøtzsche PC, Vandenbroucke JP. The Strengthening the Reporting of Observational Studies in Epidemiology (STROBE) Statement: guidelines for reporting observational studies. Int J Surg. 2014;12(12):1495–9.

22. Lyngse FP, Kirkeby CT, Denwood M, Christiansen LE, Mølbak K, Møller CH, et al. Transmission of SARS-CoV-2 Omicron VOC subvariants BA.1 and BA.2: Evidence from Danish Households. medRxiv (Preprint). 2022:2022.01.28.22270044.

23. Genomic overview of SARS-CoV-2 in Denmark: the Danish Covid-19 Genome Consortium; [cited 2022 April 8]. Available from: https://covid19genomics.dk/statistics.

24. Ioannidis JPA. Factors influencing estimated effectiveness of COVID-19 vaccines in non-randomised studies. BMJ Evidence-Based Medicine. 2022:bmjebm-2021-111901.

25. Fonager J, Bennedbæk M, Bager P, Wohlfahrt J, Ellegaard KM, Ingham AC, et al. Molecular epidemiology of the SARS-CoV-2 variant Omicron BA.2 sub-lineage in Denmark, 29 November 2021 to 2 January 2022. Euro Surveill. 2022;27(10).

26. Ioannidis JPA. Benefit of COVID-19 vaccination accounting for potential risk compensation. NPJ Vaccines. 2021;6(1):99.

27. Goldszmidt R, Petherick A, Andrade EB, Hale T, Furst R, Phillips T, et al. Protective Behaviors Against COVID-19 by Individual Vaccination Status in 12 Countries During the Pandemic. JAMA Netw Open. 2021;4(10):e2131137.

